# Role of Thromboinflammation Interaction in the Serious-Critical COVID-19 Pneumonia: A Single-center Retrospective Study

**DOI:** 10.1101/2024.04.19.24306109

**Authors:** Jiatao Li, Shimeng Liu

## Abstract

**Aim:** To describe the potential role of thromboinflammation in thromboembolism and the progress of coronavirus disease 2019 (COVID-19).

**Methods:** The retrospective study included sequentially hospitalized patients with the diagnosis of COVID-19 pneumonia during the first pandemic (December 1, 2022, to January 31, 2023) at a medical center in Beijing, China. Risks of critical medical support requirements, thrombosis events, and death were valued in the multivariate logistic regression models, where age (≥ 80 vs. < 80 years old), nadir platelets level (≤100 vs. > 100 10^9/L), C-reactive protein (CRP) level at admission (≥ 80 vs. < 80 mg/L), D-dimer level at admission (≥ 1.0 vs. < 1.0 mg/L) were dichotomized.

**Results:** A total of 88 patients were included (median age 77 years, 72% male). During hospitalization, 35 (40%) patients required critical medical support, 22 (29%) had disseminated intravascular coagulation (DIC), 11 (13%) had radiologically supported thrombosis events, and 26 (30%) died. Increased age (odds ratio [OR]: 5.22,95% confidence interval [CI]: 1.27-21.53; *p*=0.02), elevated CRP levels (OR: 9.26, 95% CI: 2.24-38.37; *p*<0.01), decreased nadir platelet level (OR: 13.47, 95% CI: 1.91-94.84; *p*<0.01), elevated D-dimer level (OR: 5.21, 95 CI%: 0.96-28.21; *p*=0.06) were associated with the requirements of critical care. Increased age (OR: 10.87, 95% CI: 3.05-38.80; *p*<0.01), elevated CRP levels (OR: 6.14, 95% CI:1.68-22.47; *p*<0.01), decreased nadir platelet levels (OR: 5.96, 95% CI: 1.34-26.47; *p*=0.02) and D-dimers (≥1.0 vs. <1.0 mg/L, OR: 2.26, 95% CI: 0.68-7.44; *p*=0.18) were associated with death. Increased age (OR: 2.40,95% CI: 0.60-9.67; *p*=0.22), decreased nadir platelet levels (OR: 1.31, 95% CI: 0.22-7.96; *p*=0.77), inflmmation status, including D-dimers (≥1.0 vs. <1.0 mg/L, OR:4.62, 95% CI: 0.89-24.04; *p*=0.07) and CRP (≥80 vs. <80 mg/mL, OR:5.44, 95% CI: 1.05-28.27; *p*=0.04) were significantly associated with the risks of thromboembolism.

**Conclusions:** The retrospective study indicated thromboinflammation promoted thromboembolism and was associated with the outcomes of COVID-19 pneumonia.

## Introduction

Since the first case of the coronavirus disease 2019 (COVID-19) was diagnosed in the city of Wuhan,China, the disease became the global pandemic disease. The new human coronavirus SARS coronavirus 2 (SARS-CoV-2) was the cause of the COVID-19,^1^ Around 80% of COVID-19 were asymptomatic or mildly symptomatic, but about 10% developed severe respiratory symptoms.^2^

COVID-19 causes a spectrum of diseases, where patients can develop a severe inflammatory state.^3^ Severe COVID-19 sepsis is associated with an inflammatory storm, with increased inflammatory markers, including C-reactive protein (CRP), ferritin, fibrin degradation products and D-dimer, et al.^4^ Interleukin (IL)-1, IL-6, and tumor necrosis factor could trigger acute endothelial cell activation.^5^ Damage to blood vessel walls or inflammatory triggers can lead to thromboinflammation, which is a process that links inflammation and thrombosis.^6^ The interaction between leukocytes and cytokines might contribute to platelet activation; meanwhile, platelets can also lead to inflammatory vasculopathy and thrombosis during thromboinflammation and infection.^6^

Autopsy studies form COVID-19 found pulmonary endothelial viral inclusions and apoptosis, perivascular T-cell infiltration, increased capillary microthrombi, and increased angiogenesis.^7,8^ Local endothelial cell dysfunctions in the pulmonary microvasculature lead to COVID-19 vasculopathy and acute respiratory distress syndrome.^4^ Higher rates of multivessel thrombosis, lower primary percutaneous coronary intervention success rates, elevated D-dimer and CRP were found in the patients with ST-elevation myocardial infarction with COVID-19 infection.^9^

The unique inflammatory storm triggered by COVID-19 leads to the thromboinflammation. While the impacts of the thromboinflammation interaction in the hemostasis status, and the disease progress remain unclear.

## Materials and methods

### Patients and data collection

Sequentially hospitalized patients with COVID-19 pneumonia during the first pandemic (from December 1, 2022, to January 31, 2023) at one medical center in Beijing, China were included. Patients with mild symptoms and patients admitted for extrapulmonary conditions were excluded. All patients were confirmed with COVID-19, which was defined as the positive SARS-CoV-2 reverse-transcriptase polymerase chain reaction (RT-PCR) tests by nasopharyngeal/oropharyngeal swabs. This study was approved by the Institutional Review Board. Informed consents were provided to the family relatives or patients. Data collected included demographics, relevant comorbidities, endotracheal intubation, length of hospital stay, completion of hospitalization (hospital discharge or death at hospital), arterial/venous thrombotic events, medications, and laboratory parameters.

### Diagnosis of COVID-19

All patients were diagnosed with the SARS-CoV-2 infection using RT-PCR for detection of viral Ribonucleic Acid (RNA) from nasopharyngeal/oropharyngeal swabs. The classification of the severity of SARS-CoV-2 infection was defined as previously described:^10^ mild disease included patients with non-pneumonia or mild pneumonia, severe disease included patients with dyspnea and hypoxia requiring supplementary oxygen, and critical disease included patients with respiratory failure requiring assisted ventilation, septic shock, and/or multi-organ dysfunctions.

Critical illness was defined as the requirements for endotracheal intubation and mechanical ventilation; the patients for whom endotracheal intubation was clinically indicated but who chose to forego it (those with a “do not rescue” status) were also included as critical illness.

### Thrombotic and disseminated intravascular coagulation (DIC) events

The incidence of thrombotic events in COVID-19 patients was assessed. The composite outcome consisted of acute pulmonary embolism (PE), deep-vein thrombosis (DVT), ischemic stroke, myocardial infarction, or systemic arterial embolism. Diagnostic examinations were applied only if thrombotic complications were clinically suspected.

DIC was evaluated with the clinical assessment combined with laboratory parameters, as defined by the International Society for Thrombosis and hemostasias (ISTH);^11^ The ISTH score relies on platelet count, level of fibrin markers (including D-dimer), prolonged prothrombin time (PT), and fibrinogen level.

PE and DVT were confirmed radiographically. Myocardial infarction was diagnosed with the clinical criteria together with biomarker elevation and electrocardiographic changes. Ischemic stroke was diagnosed with neurological defects and brain imaging supports. Synchronously diagnosed DVT, PE, or arterial thrombosis in the same patient were considered one thrombosis event.

### Classification of thrombocytopenia

Thrombocytopenia was classified according to the nadir platelet count during the hospital stay: mild (100-149 10^9/L), moderate (50-99 10^9/L), and severe (< 50 10^9/L). We classified each category of developed thrombocytopenia as an incident if the platelet count was above one of these three defined thresholds upon arrival to the emergency room and subsequently decreased to one of the categories previously defined at any time during the hospital stay.

### Statistical analysis

Continuous variables were described as median (interquartile range [IQR]) if they were not normally distributed, while categorical variables were described as number (percentage). We used χ^*2*^ tests or Fisher’s exact tests to examine the distributions of categorical variable, while Kruskal-Wallis tests were used to examine the distributions of the medians (for the non-normally distributed numerical variables), Multivariable logistic regression models were used to evaluate whether thrombosis or inflammatory parameters were associated with critical illness, death, and thrombosis events. Thresholds used in models for each biomarker were chosen based on a combination of biological relevance and the distribution of the data. Logistic regression model evaluating used the stepwise methods to select variables with *p* < 0.001 in univariate analyses. All statistical analyses were performed using SAS version 9.4. The threshold for statistical significance was *p* < 0.05.

### Patient and Public Involvement

None.

## Results

The study included 88 patients with proven serious-critical COVID-19 pneumonia admitted between December 2022 to January 2023 (Figure 1). To note, the whole patients had the first-ever COVID-19. All patients received at least standard doses of thromboprophylaxis except under contradictions.

**Figure 1:**
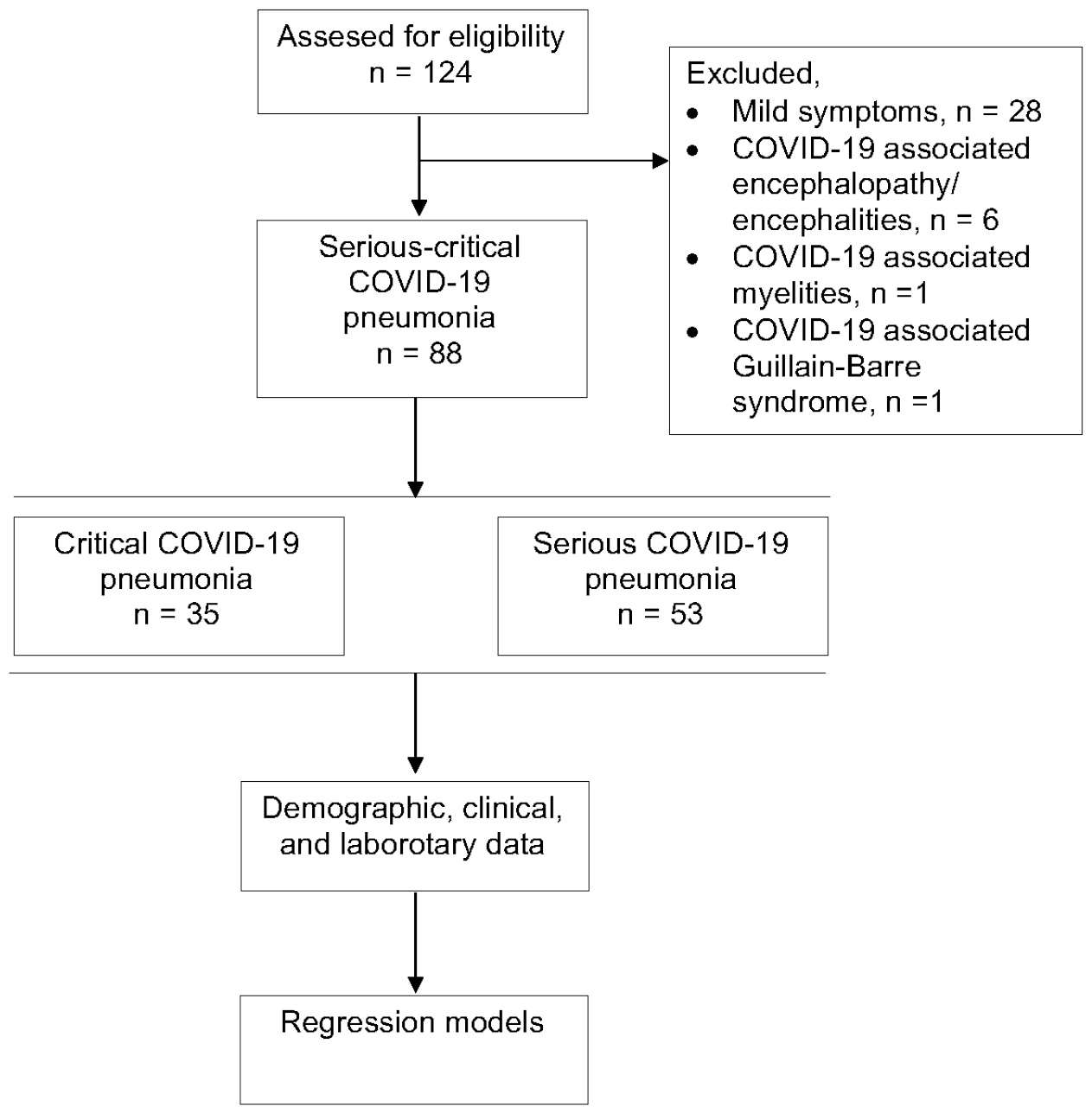
Data flow of patient selection. From December 2022 to January 2023, a total of 124 patients with proven COVID-19 were admitted. Patients with mild symptoms (n=28), patients with COVID-19 associated encephalopathy/encephalitis (n=6), 1 patient with COVID-19 associated myelitis and 1 patient with COVID-19 associated Guillain-Barre syndrome were excluded from the data analysis. Finally, 88 patients with proven serious-critical COVID-19 pneumonia were included in the study.

### Baseline characteristics

A total of 88 patients were included (median age 77 years; 72% male) (Table 1). Thiry-five (40%) patients required critical medical support. Patients who required critical medical support were elder (median age: 81 vs. 73 years; *p*<0.01). The most common comorbidities were diabetes (32; 36%), cardiovascular disease (19; 22%), and cancer 18 (20%). We found a significant disparity in having a history of anemia (serious vs. critical illness: 0 vs. 11%; *p*=0.02). At triage, 84% of patients were febrile, 25% had shortness of breath. Patients will critical pneumonia were admitted at 7 days of symptom onset, which was shorter than that of patients with serious illness (10 days) (*p*<0.01).

**Table 1.** Baseline characteristics.

|  | Whole (n=88) | Critical (n=35) | Serious (n=53) | *p* value |
| --- | --- | --- | --- | --- |
| Age (years) | 77 (68-83) | 81 (76-86) | 73 (65-78) | < 0.001 |
| Sex (M) | 63 (72) | 23 (65) | 40 (75) | 0.323 |
| Comorbidities |  |  |  |  |
| Diabetes | 32 (36) | 14 (40) | 18 (34) | 0.564 |
| Cardiac vascular disease | 19 (22) | 7 (20) | 12 (23) | 0.767 |
| Cancer | 18 (20) | 7 (20) | 11 (21) | 0.932 |
| Stroke | 15 (17) | 4 (11) | 11 (21) | 0.255 |
| Pre-respiratory disease | 14 (16) | 7 (20) | 7 (13) | 0.394 |
| Peripheral artery disease | 9 (10) | 4 (11) | 5 (9) | 1.000 |
| Hypothyroidism | 8 (9) | 5 (14) | 3 (6) | 0.257 |
| Heart failure | 6 (7) | 3 (9) | 3 (6) | 0.679 |
| Neurological degeneration | 6 (7) | 5 (14) | 1 (2) | 0.034 |
| Renal failure | 5 (6) | 3 (9) | 2 (4) | 0.383 |
| Anemia | 4 (5) | 4 (11) | 0 | 0.023 |
| Atrial fibrillation | 3 (3) | 2 (6) | 1 (2) | 0.561 |
| Symptoms onset (days) | 10 (7-15) | 7 (3-10) | 10 (10-17) | 0.001 |
| Fever | 74 (84) | 28 (80) | 46 (88) | 0.394 |
| Highest Temperature (°C) | 38.5 (38.0-39.0) | 38.3 (38.0-39.0) | 38.5 (38.0-39.0) | 0.431 |
| Cough/Sputum production | 82 (93) | 32 (91) | 50 (94) | 0.679 |
| Short of Breath | 22 (25) | 12 (34) | 10 (19) | 0.102 |

Data are presented as n (%) or median (interquartile range). M: male. *p* value, comparison between patients with critical and serious pneumonia.

### Laboratory results

The distribution of median CRP (134.88 vs. 52.54 mg/L; *p*<0.01), white blood cell (WBC) counts (9.58 vs. 7.44 10^9/L; *p*=0.03), lymphocyte counts (0.55 vs. 0.90 10^9/L; *p*<0.01), and platelet counts (163 vs. 189 10^9/L; *p*=0.01) were significantly different whether the patients required critical care supports or not (Table 2). To note, the nadir platelet counts during hospital stay were lower (135 vs. 173 10^9/L; *p*=0.01), and more patients underwent thrombocytopenia (29 vs. 19 %; *p*=0.02) in the patients with critical conditions (Table 2). Patients in critical conditions had higher neutrophil-to-lymphocyte ratio (NLR) (12.40 vs. 6.58; *p*<0.01).

**Table 2.** The laboratory results of the patients.

|  | Whole (n=88) | Critical (n=35) | Serious (n=53) | *p* value |
| --- | --- | --- | --- | --- |
| Blood routine tests |  |  |  |  |
| CRP ( < 10.00 mg/L) | 78.09 (38.32-141.38) | 134.88 (72.10-185.01) | 52.54 (14.78-110.00) | <0.001 |
| WBC (3.5-9.5 10^9/L) | 7.83 (5.52-10.70) | 9.58 (6.16-13.95) | 7.44 (5.06-9.30) | 0.032 |
| Lymphocyte (1.1-3.2 10^9/L) | 0.74 (0.46-1.20) | 0.55 (0.36-0.76) | 0.90 (0.54-1.30) | 0.003 |
| Neutrophil (1.8-6.3 10^9/L) | 6.48 (3.98-8.78) | 8.48 (5.25-12.59) | 5.75 (3.75-7.48) | 0.004 |
| NLR | 8.62 (4.90-15.77) | 12.40 (6.50-22.98) | 6.58 (4.10-9.38) | <0.001 |
| RBC (125-350 10^9/L) | 4.03 (3.61-4.54) | 3.77 (3.22-4.31) | 4.20 (3.87-4.64) | 0.004 |
| Hb (115-150 g/L) | 128 (116-143) | 121 (98-129) | 132 (121-145) | 0.008 |
| PLT (125-350 10^9/L) | 185 (141-264) | 163 (105-209) | 189 (164-297) | 0.011 |
| Nadir PLT | 156 (115-193) | 135 (76-185) | 173 (136-235) | 0.001 |
| Thrombocytopenia |  |  |  |  |
| PLT: ≥ 150 10^9/L | 68 (77%) | 25 (71%) | 43 (81%) | 0.023 |
| PLT: 100-149 10^9/L | 12 (14%) | 3 (9%) | 9 (17%) |  |
| PLT: 50-99 10^9/L | 5 (6%) | 4 (11%) | 1 (2%) |  |
| PLT: < 50 10^9/L | 3(3%) | 3 (9%) | 0 |  |
| Coagulation board |  |  |  |  |
| FDP (0-5 µg/mL) | 4.27 (2.25-8.14) | 5.37 (3.75-15.56) | 2.80 (1.77-5.59) | 0.001 |
| D-Dimer (0-1.5 µg/mL) | 1.43 (0.77-5.10) | 2.69 (1.25-8.96) | 1.10 (0.60-3.05) | 0.002 |
| PT (9.40-12.50 sec) | 12.35 (11.50-13.80) | 12.85 (11.80-14.00) | 12.05 (11.30-13.20) | 0.042 |
| Fibrinogen (2.00-4.00 g/L) | 4.51 (3.92-6.09) | 5.47 (3.97-6.11) | 4.44 (3.92-6.00) | 0.430 |
| Cardiacmarkers |  |  |  |  |
| BNP (0-100 pg/ml) | 112.9 (28.1-350.0) | 194.8 (87.8-671.0) | 62.6 (19.6-221.0) | 0.002 |
| CK-MB (0-5.00 ng/ml) | 3.12 (1.59-6.10) | 3.59 (2.14-7.97) | 2.51 (1.19-5.00) | 0.022 |
| TnI (0-0.034 ng/ml) | 0.014 (0.006-0.065) | 0.049 (0.012-0.210) | 0.007 (0.003-0.031) | <0.001 |
| Serum biochemical tests |  |  |  |  |
| K+ (3.50-5.30 mmol/L) | 4.02 (3.69-4.40) | 3.85 (3.59-4.30) | 4.07 (3.80-4.40) | 0.169 |
| Na+ (135.0-145.0 mmol/L) | 135.3 (132.3-138.4) | 135.0 (130.1-138.7) | 136.6 (133.0-138.0) | 0.326 |
| ALT (0.0-41.0 U/L) | 24.7 (14.5-39.3) | 19.8 (13.5-27.0) | 32.2 (17.0-42.4) | 0.017 |
| AST (0.0-42.0 U/L) | 29.9 (20.5-42.1) | 33.0 (20.0-45.0) | 28.0 (21.0-41.0) | 0.406 |
| ALB (35.0-55.0 g/L) | 32.9 (30.0-35.2) | 31.3 (29.0-33.0) | 33.8 (31.5-36.5) | 0.003 |
| Cr (31.7-93.3 µmol/L) | 78.0 (60.6-103.2) | 91.6 (63.9-175.4) | 71.0 (60.5-88.5) | 0.020 |
| Ferritin (22.0-322.0 ng/ml) | 508.2 (299.9-1223.5) | 906.5 (631.6-1328.5) | 414.0 (261.5-1035.5) | 0.142 |
| Arterial blood gas analysis |  |  |  |  |
| Sputum culture |  |  |  |  |
| Bacteria positive | 21 (29%) | 17 (52%) | 4 (10%) | <0.001 |
| Fungi positive | 7 (10%) | 5 (15%) | 2 (5%) | 0.233 |

Data are presented as n (%) or median (Interquartile range). CRP: C-reactive protein; WBC: white blood cell; NLR: neutrophil-to-lymphocyte ratio; RBC: red blood cell; Hb: hemoglobin; PLT: platelet; BNP: brain natriuretic peptide; CK-MB: creatine kinase-MB; TnI: troponin I; ALT: aminotransferase; AST: aspartate aminotransferase; ALB: albumin; Cr: creatin; FDP: fibrinogen degradation products; PT: prothrombin time; *p* value, comparison between patients with serious and critical pneumonia.

The levels of fibrinogen degradation products (FDP) (5.37 vs. 2.80 μg/mL; *p*<0.01) and D-dimers (2.69 vs. 1.10; *p*<0.01) were higher, and prothrombin time (PT) (12.85 vs. 12.05 sec; *p* =0.04) was longer in patients with critical conditions (Table 2).The cardiac markers (troponin I: 0.049 vs. 0.007; *p*<0.01) and serum creatine (91.6 vs. 71.0 μmol/L; *p*=0.02) were higher, while serum albumin at baseline was lower (31.3 vs. 33.8 g/L; *p*<0.01) in patients who required critical care support (Table 2). The rate of respiratory bacterial co-infection was higher in patients who required critical medical supports (52% vs. 10%; *p*<0.01) (Table 2).

### The medical data and outcomes

During hospitalization, 27 (31%) patients received invasive mechanical ventilation, 24 (28%) received Nirmatrelvir-Ritonavir, 70 (80%) received glucocorticoid and 57 (65%) received anti-coagulation (Table 3).

**Table 3.**
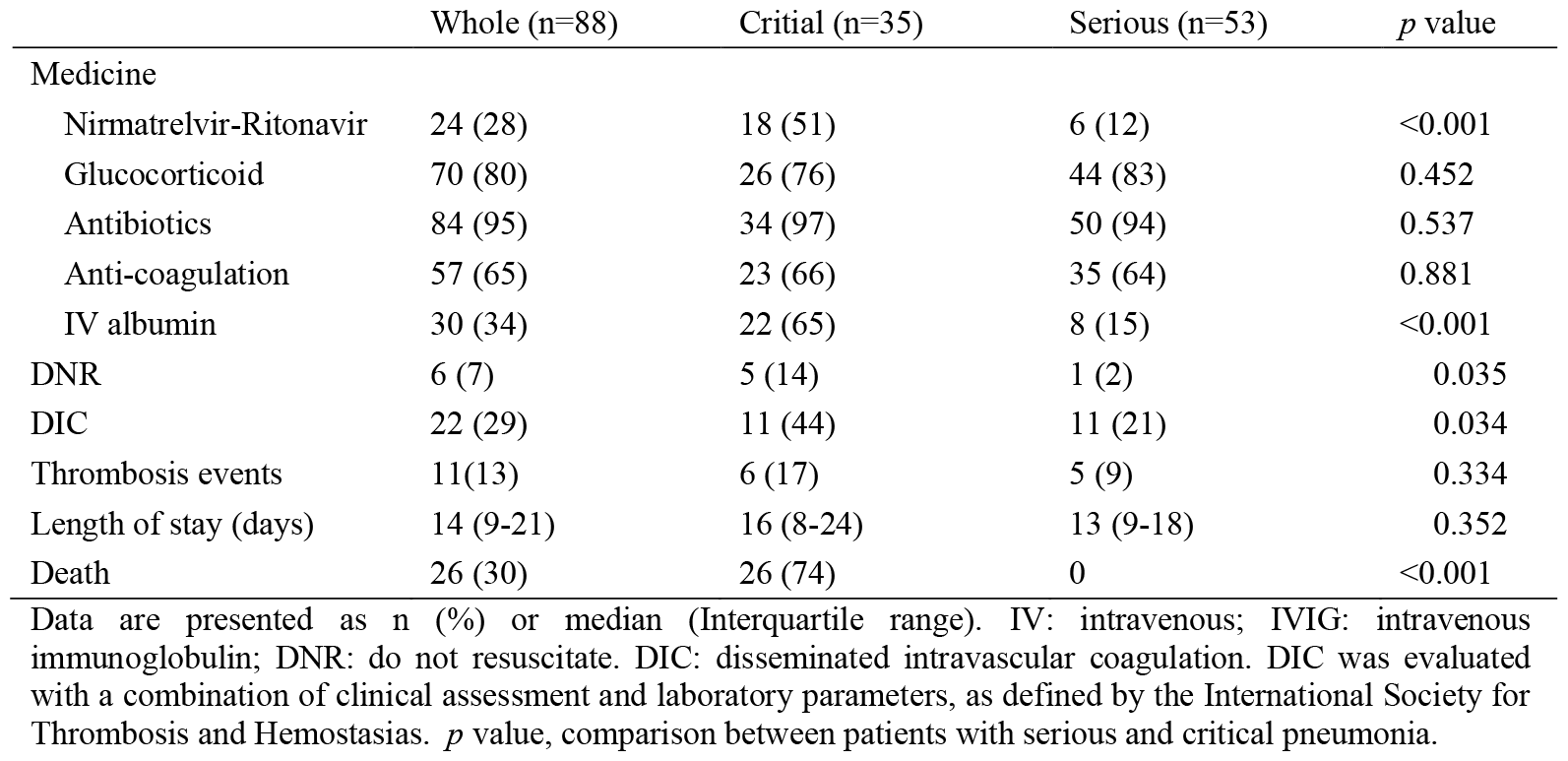
The medical data and outcomes of patients.

|  | Whole (n=88) | Critical (n=35) | Serious (n=53) | *p* value |
| --- | --- | --- | --- | --- |
| Medicine |  |  |  |  |
| Nirmatrelvir-Ritonavir | 24 (28) | 18 (51) | 6 (12) | <0.001 |
| Glucocorticoid | 70 (80) | 26 (76) | 44 (83) | 0.452 |
| Antibiotics | 84 (95) | 34 (97) | 50 (94) | 0.537 |
| Anti-coagulation | 57 (65) | 23 (66) | 35 (64) | 0.881 |
| IV albumin | 30 (34) | 22 (65) | 8 (15) | <0.001 |
| DNR | 6 (7) | 5 (14) | 1 (2) | 0.035 |
| DIC | 22 (29) | 11 (44) | 11 (21) | 0.034 |
| Thrombosis events | 11(13) | 6 (17) | 5 (9) | 0.334 |
| Length of stay (days) | 14 (9-21) | 16 (8-24) | 13 (9-18) | 0.352 |
| Death | 26 (30) | 26 (74) | 0 | <0.001 |

Data are presented as n (%) or median (Interquartile range). IV: intravenous; IVIG: intravenous immunoglobulin; DNR: do not resuscitate. DIC: disseminated intravascular coagulation. DIC was evaluated with a combination of clinical assessment and laboratory parameters, as defined by the International Society for Thrombosis and Hemostasias. *p* value, comparison between patients with serious and critical pneumonia.

The incidence of DIC was higher in the patients with critical conditions (44 vs. 21%; *p*=0.03); while the incidence of thrombosis was similar between the groups (17% vs. 9%; *p*=0.33). Twenty-six (74%) patients died in critical condition, but none of the patients with serious pneumonia died (Table 3).

### The impact of thromboinflammation

In the multivariate logistic regression models (Figure 2), increased age (odds ratio [OR]: 5.22,95% confidence interval [CI]: 1.27-21.53; *p*=0.02), elevated CRP levels (OR: 9.26, 95% CI: 2.24-38.37; *p*<0.01), decreased nadir platelet level (OR: 13.47, 95% CI: 1.91-94.84; *p*<0.01), elevated D-dimer level (OR: 5.21, 95 CI%: 0.96-28.21; *p*=0.06) were associated with the requirements of critical care. Increased age (OR: 10.87, 95% CI: 3.05-38.80; *p*<0.01), elevated CRP levels (OR: 6.14, 95% CI:1.68-22.47; *p*<0.01), decreased nadir platelet levels (OR: 5.96, 95% CI: 1.34-26.47; *p*=0.02) and D-dimers (≥1.0 vs. <1.0 mg/L, OR: 2.26, 95% CI: 0.68-7.44; *p*=0.18) were associated with death. Increased age (OR: 2.40,95% CI: 0.60-9.67; *p*=0.22), decreased nadir platelet levels (OR: 1.31, 95% CI: 0.22-7.96; *p*=0.77), inflmmation status, including D-dimers (≥1.0 vs. <1.0 mg/L, OR:4.62, 95% CI: 0.89-24.04; *p*=0.07) and CRP (≥80 vs. <80 mg/mL, OR:5.44, 95% CI: 1.05-28.27; *p*=0.04) were significantly associated with the risks of thromboembolism.

**Figure 2:**
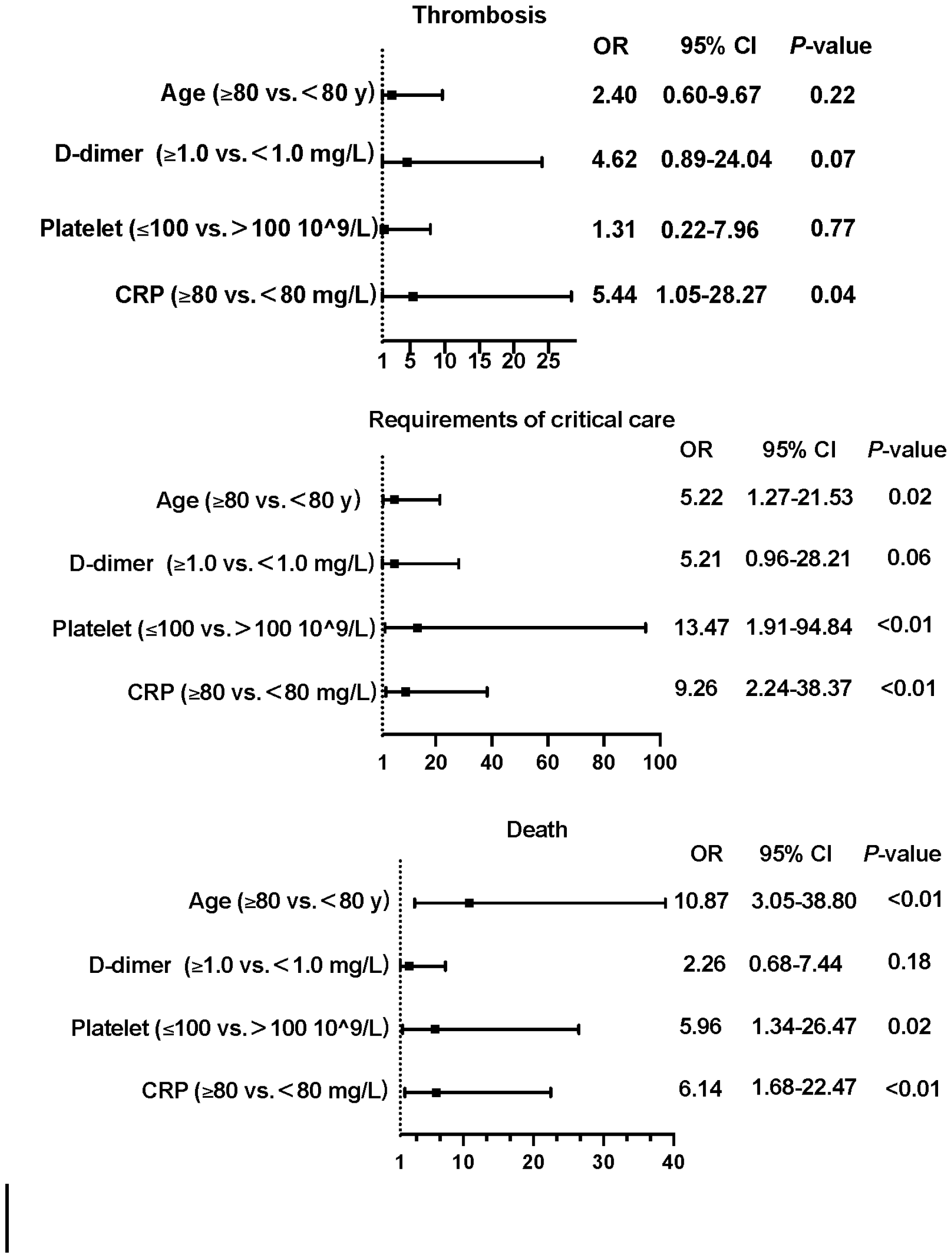
Logistic regression models. Increased age (odds ratio [OR]: 5.22,95% confidence interval [CI]: 1.27-21.53; *p*=0.02), elevated CRP levels (OR: 9.26, 95% CI: 2.24-38.37; *p*<0.01), decreased nadir platelet level (OR: 13.47, 95% CI: 1.91-94.84; *p*<0.01), elevated D-dimer level (OR: 5.21, 95 CI%: 0.96-28.21; *p*=0.06) were associated with the requirements of critical care. Increased age (OR: 10.87, 95% CI: 3.05-38.80; *p*<0.01), elevated CRP levels (OR: 6.14, 95% CI:1.68-22.47; *p*<0.01), decreased nadir platelet levels (OR: 5.96, 95% CI: 1.34-26.47; *p*=0.02) and D-dimers (≥1.0 vs. <1.0 mg/L, OR: 2.26, 95% CI: 0.68-7.44; *p*=0.18) were associated with death. Increased age (OR: 2.40,95% CI: 0.60-9.67; *p*=0.22), decreased nadir platelet levels (OR: 1.31, 95% CI: 0.22-7.96; *p*=0.77), inflmmation status, including D-dimers (≥1.0 vs. <1.0 mg/L, OR:4.62, 95% CI: 0.89-24.04; *p*=0.07) and CRP (≥80 vs. <80 mg/mL, OR:5.44, 95% CI: 1.05-28.27; *p*=0.04) were significantly associated with the risks of thromboembolism.

## Discussion

We described the potential role of thromboinflammation interaction in thromboembolism and the progress of COVID-19 pneumonia in the hospitalized patients at one tertiary hospital from December 2022 to January 2023, when the COVID-19 outbreak occurred for the first time in city. To mention, the whole study patients had the first-ever SARS-CoV2 infections. Inflammation and thrombosis are the hallmark features of COVID-19. Significant elevated inflammatory biomarkers (CRP, D-dimers) and thrombocytopenia were shown in patients who required critical care support and died. Thromboembolism was associated with DIC and inflammation, but not the disease severity or patients’ age.

We found advanced age, elevated CRP, D-dimers, decreased platelets were associated with critically ill and death. The study was consistent with a retrospective analysis of 1449 Chinese patients, age (OR: 1.18, 95% CI: 1.02-1.36]) and baseline D-dimer (OR: 3.18; 95% CI: 1.48-6.82), fibrinogen (OR: 6.45. 95% CI: 1.31-31.69), platelets (OR: 0.95; 95% CI: 0.90-0.99), CRP (OR: 1.09. 95% CI: 1.01-1.18), and lactate dehydrogenase (OR:1.03, 95% CI:1.01-1.06]) correlated with an increased risk of death.^12^

The NLR, calculated as a ratio between the neutrophil and lymphocyte counts measured in peripheral blood, was increased in the patients with critical conditions. In the SARS-CoV-2 infections, neutrophil accumulation was impaired by the inhibition of interferon-γ, leading to an increase in the neutrophils;^13^ the reduction of lymphocyte was another characteristic and the rate of this reduction inversely correlates with the disease severity,^14^ which might due to the functional exhaustion of cytotoxic lymphocytes.^15^

The other key early laboratory observations were elevated plasma D-dimer concentrations and CRP, reflecting the inflammatory response. The marked elevation of D-dimers could be led by coagulation activation from viremia and cytokine storm, meanwhile superinfection and organ dysfunction are other possible causes.^3^ As the reports, D-dimers and FDP progressively increased with COVID-19 severity, but the fibrinogen level stayed elevated,^16-18^ which was in contrast to DIC. The coagulopathy under the COVID-19 condition is most likely due to the inflammatory response and the endothelial activation/damage,^19^ with a prominent elevation of fibrinogen and D-dimer/fibrinogen degradation products.^3^

Thrombocytopenia, common in acute infections and can correlate with disease severity,^20,21^ was noticeable in our study as well. In critically ill patients, thrombocytopenia was associated with worse outcomes, the prevalence of which was 8-56 %.^22^ The causes of thrombocytopenia included sepsis, DIC, consumption, heparin-induced thrombocytopenia, hemophagocytic syndrome, et al,.^23^

COVID-19 was reported to predispose to both venous and arterial thromboembolism.^24,25^ Despite systematic thrombosis prophylaxis, the incidence of thrombotic complications was 13% in whole patients, and 17% in patients with critical illness. The incidence rates of thrombotic complications were lower than in prior report, where the cumulative incidence of venous and arterial thrombotic events was around 30%.^24^ At our center, computed tomography pulmonary angiography was not required for screening PE, as to avoid radiation exposure and risk of contrast medium–induced nephropathy.^26^ If VTE screenings had been applied, the incidence of thrombosis might have been higher. Elevated D-dimer at initial presentation, platelet count >450 10^9^/, CRP >100 mg/L and erythrocyte sedimentation rate (ESR) >40 mm/h predict the thrombosis during hospitalization. DIC is a systemic activation of coagulation, which can be caused by severe infections.^27^ The 29% incidence of DIC events was higher than in prior studies.^28^ We found a significant association between DIC/CRP with the thromboembolism as well. Elevated cardiac markers, including brain natriuretic peptide and troponin T, were also noticed. The elevated cardiac markers may reflect the emergent ventricular stress induced by pulmonary hypertension.^29^

The study has limitations. First, we were lacking in further study to investigate the underlying mechanism in thromboinflammation interaction. Second, the longitudinal effects of thromboinflammation interaction over time were not evaluated. Thirdly, the sample size was small, so we did not have enough power to test the relationships between the inflammation and thrombosis progress.

The retrospective study implied the potential role of thromboinflammation in the progress of COVID-19 pneumonia. The unique inflammation storm in COVID-19 might triggered the thrombosis, which played an important role in the disease progress and thromembolism. Further studies were required to investigate the underlying mechanisms of thromboinflammattion interaction.

## Data availability statement

The data that support the findings of this study are available from the corresponding author upon reasonable request.

## Disclosure

The study did not include any relevant commercial, financial, or non-financial associations.

## Funding

This work was supported by the National Natural Science Foundation of China under Grant [82001242].

## Acknowledgements

None.

## Author Statement

Li, J: data collection and manuscript preparation; Liu, S: study design, data analysis, and manuscript improvements

## Notes

### Competing Interest Statement

The authors have declared no competing interest.

### Author Declarations

The study was approved by the IRB of Beijing Tiantan Hospital, Capital Medical University.

